# The contribution of parental health to the subsequent social assistance entry of the family with children– a nationwide register-linked birth cohort study in Finland

**DOI:** 10.1101/2023.01.20.23284822

**Authors:** Aapo Hiilamo, Markus Keski-Säntti, Sami Räsänen, Mika Niemelä, Tea Lallukka, Tiina Ristikari

## Abstract

**Aim:** Our aim in this paper is to estimate the contribution of different parental specialised health care diagnoses to the subsequent risk of entry into the social assistance system for families with children in the period 1998–2013.

**Methods:** We used longitudinal population-level register data consisting of all children born in 1997 in Finland and their registered parents (54 960 one and two-parent families with 801 336 observations in the period 1998–2013). Diagnoses assigned in public specialised health care and social assistance records were derived from nationwide administrative registers. Measures of parental socioeconomic status and previous diagnoses and the birth weight of the child were adjusted for in regression models which estimated the association between parental diagnoses and entry into the social assistance system in the following year.

**Results:** Families with a parent somatic diagnosis had a risk ratio of 1.4 for social assistance entry in the subsequent year of the diagnosis though substantial variation by diagnosis category was detected. Parent psychiatric diagnoses were linked to a higher, 3.01-fold risk of social assistance entry. Covariate adjustment reduced these risk ratios to 1.2 and 2.1, respectively. Some 2.9 percent of all social assistance entries may be attributed to parental psychiatric diagnoses while somatic health records account for another 7.2 percent, making their total contribution over a tenth of all cases.

**Conclusion:** Parental specialised health care records was associated with a higher risk of social assistance need as more interventions to support financial management are required for parents with psychiatric diagnoses.

**What is already known on this topic:**

- Disruptive life events, such as work disability, unemployment or divorce, are associated with social assistance claims and usage in the general population

**What this study adds:**

- Parents’ diagnoses, particularly when due to psychiatric conditions, are associated with a higher risk of entry into the social assistance system

**How this study might affect research, practice or policy:**

- The findings are important in enabling us to recognise target groups and in designing interventions that aim to prevent families with children from falling into economic hardship

## Background

Social assistance is the public assistance measure of last resort, when savings and income from employment and other social security benefits, such as unemployment insurance, disability insurance or housing allowance, are not available or deemed inadequate to provide an acceptable standard of living (1). The use of social assistance in families is an important proxy for child poverty (2) because, in the Nordic countries, social assistance is granted only to households with no spare income or savings to cover their necessities (1,3). Measures of child poverty and economic hardship are associated with a range of immediate, intermediate and long-term mental and physical health outcomes of the children affected (4–6), making the reduction of child poverty an important public health goal (7). In order to find sustainable ways to reduce child poverty, it is imperative to identify the key attributable factors in relation to social assistance entry. Being strictly means-tested, the use of social assistance reflects both administratively and subjectively assessed poverty (2) with around one-tenth of families with children using social assistance at least once a year (8). In Finland, the country in focus here, social assistance is granted at the individual or family level on a monthly basis as temporary relief to guarantee the constitutional right to acceptable living standards for those unable to provide for themselves.

Parental health issues may be linked to social assistance needs via reduced work ability, treatment costs and difficulties in managing financial matters but also to stigma. It is important to note that social assistance is not intended as a programme to insure against ill health. Separate public work disability insurance and medical insurance instruments exist in Finland. Nevertheless, it is well-established that disruptive life events, such as work disability, unemployment or divorce, are associated with social assistance claims and usage in the general population (2,9–12). A smaller body of studies indicates that health-related variables predict social assistance outcomes. A previous Finnish register study on social assistant recipients found that healthcare use predicted social assistance receipt in the following month (13). A Swedish study reported that persons with previous long sickness absence episodes were less likely to exit from social assistance (12). In a longitudinal Norwegian study on young adults, disability and rehabilitation increased the likelihood of social assistance entry (9).

Until now, however, there was no evidence on social assistance usage with families with children as the unit of analysis. This is an important gap because findings from the general population may not generalise to families with children. Social assistance use in families with children is also an extra policy-relevant issue because social assistance use is often intergenerational, that is, parental social assistance use predicts children’s social assistance use later in their life (14). In this study, we estimate the contribution of parental health, indicated by diagnoses assigned in specialised health care, on the subsequent risk of entry into the social assistance system. In addition to individual-level risk ratios, we also focus on population-level attributable factors of social assistance entry, that is the most important diagnoses leading to social assistance entry. This is to help policymakers and practitioners to better understand the most common causes of social assistance entry which in turn will help them to design the most optimally targeted interventions (15).

## Methods

We used prospective register-based birth cohort data on all families with children born in Finland in 1997. This dataset was formed by combining several national administrative databases. The complete list of these databases is available elsewhere (16). A unique personal identification number assigned to all legal residents allows reliable matching of administrative data sources (17). Data on registered parents and children were linked using the Population Information System of Finland. The registers used are of the highest quality and are regularly checked for potential errors and omissions. The cohort is maintained by the Finnish Institute for Health and Welfare and approved by the ethical committee of the institute (decision § 5/2013). The data are analysed for public interest and scientific purposes. Being a register-based cohort, the members of the study were not contacted for informed consent. The data is analysed in line with the regulations. Permission to analyse the linked administrative data was obtained from the registrars.

The study population for this study is all families with children born in Finland in 1997. We use information from the child to define the study population as the cohort study is constructed around the children born in 1997. The original cohort consisted of Finnish Birth Cohort, consisting of all children born alive in 1997 (58 802 children) but, as our unit of analysis is the family, we exclude children with missing parental data (641), children who moved abroad (1 255) and other siblings of the same mother born in 1997 (885). Finally, we excluded families in which children never lived with their parents as well as families that received social assistance every year between 1998 and 2013 (in this way, a further1061 were further excluded). We ended up with 54 960 families. For these families, we constructed yearly data spanning the period 1998-2013 and then included observations in which the parents lived with the children and in which the family did not receive social assistance at year *t* because our focus is on social assistance entries. Supplementary figure 1 shows this sample selection process. Thus, our analysis consists of 801 336 observations for unadjusted models. These observations represent the years in which the families did not receive any social assistance. For each year, the data consists of measures of families’ socioeconomic circumstances, health and education. Our exposure (health care records) and outcome (social assistance) are derived using parents’ information.

### Outcome

Our outcome variable is social assistance entry. The Finnish social assistance system consists of three types of means-tested assistance: basic, supplementary and preventative social assistance. In this study we focus on basic and supplementary social assistance, two benefits that are termed primary social assistance (18). All persons and families residing in Finland with little income or assets, are eligible for primary social assistance. During the study period, primary social assistance was granted by local municipalities. Primary social assistance covers some of the basic expenses of living. For example, for a single adult household, social assistance was some 500 euros / month in 2019. In addition, on a means-tested basis, housing costs, medical expenses and some other necessities are covered. While intended as a short-term last-resort relief to overcome temporary economic difficulties, social assistance periods are not typically short, with an average of 6 months (19). Some 70 percent of people receiving social assistance are unemployed and only five percent of social assistance recipients have some income from employment (20).

Social assistance entry was ascertained from the Register of Social Assistance which contains all households receiving social assistance in the country. The data are originally obtained from municipalities. The registrar is the Finnish Institute for Health and Welfare. In this study we define social assistance entry as when a parent of the family is a recipient of social assistance at least once within the year *t*+1. As the data is restricted to families receiving no social assistance use in year *t*, this reflects social assistance entry. If a parent is not living with their children (i.e., a divorced parent), then this parent’s social assistance record is not taken into account.

### Exposure

Our exposure variables are parental diagnoses in specialised health care. We conceptualise, in line with previous work (13), specialised healthcare diagnoses as proxy variables for parental health. We define an exposed family as one with a parent who had at least one specialised health care record with the given primary diagnosis within the given calendar year *t*, that is, the last year before potential entry to social assistance system..We do not take into consideration secondary diagnoses). As with social assistance usage, when one of the parents lived at a different address than their children, we ignore this parent’s exposure. It is worth noting that we have no data on the health care records of step-parents of the children.

Data on public specialised healthcare diagnoses were derived from the national Care Register (Hilmo) (21). Finland has a universal health care system with low user charges. Data consist of all specialised inpatient and outpatient care visits to public hospitals in the country. The data do not contain private or occupational health care visits nor primary care episodes. Private health care is a minor sector in the country and occupational health care visits are typically confined to a basic care level. We discuss the limitations regarding primary care visits in the discussion section.

We repeat the analysis using more detailed diagnoses as the main diagnostic categories associated with a particular medical specialty of the Finnish ICD-10 manual. In addition to the diagnosis categories, we repeat the analysis using parental unemployment as an alternative exposure variable. This is to provide a tangible point of comparison. We also created a combined somatic diagnoses category which included all diagnoses, except O, F, and Z -categories.

### Covariates

We adjust our models for covariates ascertained from nationwide administrative registers. Our selection of the covariates was guided by the previous evidence on the common socioeconomic risk factors of ill-health (22–25) and data availability as register data used only include a limited set of potential covariates. Covariates included the birth weight, sex and month of the child born in 1997 and several parental characteristics in 1997, measured separately for registered mothers and fathers: personal income, diagnosis, education, living arrangement and unemployment. We also included a dichotomous variable measuring past social assistance use. The birth weight of the child was derived from the national birth register. Even though the analysed families may have other children, we use the birth weight of children born in 1997 as an important proxy of families’ living situation. This is because birth weight reflects both parental health and socioeconomic factors, both of which contributes to the risk of specialised health care records. The personal incomes of the registered father and mother, measured separately, are taken from the earnings-and accrual register of the Finnish Centre for Pensions. Personal incomes are measured yearly. Previous diagnoses are the given as a one-year lagged variable (for the lag diagnoses measured in year 1998, we do not have outpatient diagnoses data due to data availability). Parental education is taken from the Statistics Finland education register. Unemployment variables for both parents were constructed by combining information from the benefits register of the Finnish Social Insurance Institution and the earnings-and accrual register of the Finnish Centre for Pensions’ administration. These variables are constructed using data from a range of unemployment-related social benefits use. All covariates were measured a year before (year *t*-1) the exposure to avoid bias arising from controlling for mediators.

### Statistical analysis

We calculated risk ratios for social assistance entry using Poisson regression models. We chose the Poisson model for the dichotomous social assistance entry outcome to provide us with risk ratios, that is, the social assistance entry risk of the selected diagnosed group to the risk of the not diagnosed group. We fit all models separately for each diagnoses category. We do not mutually adjust different diagnoses in the models due to multimorbidity and overadjustment issues. In the adjusted model, we include time as a categorical predictor and the control variables described above. We also include time and parental income interactions to take into consideration price increases. Families with missing data on the birth weight of the child were not included in the adjusted model (1724 observations/117 families). The study design is described in Supplementary figure 2. For a causal interpretation, there should be no unmeasured confounding over and above measured variables.

In the unadjusted model, we do not report confidence intervals given that the population of interest is observed in the data. In the adjusted model, we calculated confidence intervals using sandwich estimation to correct the overly wide confidence intervals of the Poisson models for binary outcomes (26).

We then calculate population attributable fractions (PAF), that is, the proportion of social assistance entries that are attributable to a given diagnosis. We use the Miettinen formula to calculate PAFs from the proportion of a given diagnosis among the social assistance entrants (P_c) and from the risk ratio of social assistance entry (rr): p_c*(rr-1)/rr (15,27). Unadjusted PAF was calculated using unadjusted risk ratios. Adjusted PAF was calculated using adjusted risk ratios from the Poisson model described above.

## Results

There were 19,174 social assistance entries, corresponding to 2.4 percent of all observations (Table 1). Some 21.9 percent of families experienced at least one transition to social assistance during the study period. Parental somatic diagnoses were recorded in 298 074observations (37.2% of all observations) with some 95.2 percent of families experiencing a parental somatic diagnosis. Parental psychiatric F-diagnoses were recorded in 15 599 observations (1.9%) with some 12.6 percent of families experiencing a parental psychiatric diagnosis. The three most common diagnoses were factors influencing health status and contact with health services (Z00–Z99 category, recorded for some 10.9% of observations), musculoskeletal disorders (M00–M99-category, some 7.5% of observations) and symptoms, signs and abnormal clinical and laboratory findings, not elsewhere classified (R00–R99 category, some 6.3% of observations).

**Table 1.** Description of the study population. 1997 birth cohort (n_families=54 960, n_observations=801 336).

|  | Category title | incidence<br>rate | Number of<br>observations | Share (%) of families ever<br>experiencing during the study<br>period |
| --- | --- | --- | --- | --- |
| **Outcome** |  |  |  |  |
| Subsequent social<br>assistance entry<br>$t+1$ | | 2.4 | 19174 | 21.9 |
| **Predictors** |  |  |  |  |
| Any somatic father |  | 19,7 | 157577 | 78,6 |
| Any somatic<br>parents |  | 37,2 | 298074 | 95,2 |
| Any somatic<br>mother |  | 22,5 | 180044 | 82,3 |
| A00–B99 | Certain infectious and<br>parasitic diseases | 0.9 | 7200 | 10.4 |
| C00–D48 | Neoplasms | 2.7 | 21814 | 18.7 |
| D50–D89 | Diseases of the blood and<br>blood-forming organs and<br>certain disorders involving<br>the immune mechanism | 0.5 | 3618 | 3.4 |
| E00–E90 | Endocrine, nutritional and<br>metabolic diseases | 1.5 | 12299 | 8.4 |
| F00–F99 | Mental and behavioural<br>disorders | 1.9 | 15599 | 12.6 |
| G00–G99 | Diseases of the nervous<br>system | 3.1 | 25037 | 21.4 |
| H00–H59 | Diseases of the eye and adnexa | 2 | 15860 | 16.4 |
| H60–H95 | Diseases of the ear and mastoid process | 1.2 | 9655 | 9.6 |
| I00–I99 | Diseases of the circulatory system | 3.3 | 26366 | 25.1 |
| J00–J99 | Diseases of the respiratory system | 3.4 | 26955 | 27.4 |
| K00–K93 | Diseases of the digestive system | 4.6 | 36979 | 33.8 |
| L00–L99 | Diseases of the skin and subcutaneous tissue | 1.8 | 14242 | 15.7 |
| M00–M99 | Diseases of the musculoskeletal system and connective tissue | 7.5 | 60102 | 45.2 |
| N00–N99 | Diseases of the genitourinary system | 4.2 | 33838 | 32.8 |
| Q00–Q99 | Congenital malformations, deformations and chromosomal abnormalities | 0.3 | 2737 | 2.8 |
| R00–R99 | Symptoms, signs and abnormal clinical and laboratory findings, not elsewhere classified | 6.3 | 50440 | 49 |
| S00–T98 | Injury, poisoning and certain other consequences of external causes | 5.3 | 42368 | 44.6 |
| Z00–Z99 | Factors influencing health status and contact with health services (not an illness but can affect health) | 10.9 | 86990 | 69.4 |

In the full sample analysis (Figure 1), before adjusting for covariates, parental somatic diagnoses were linked to social assistance entry in the subsequent year with a risk ratio of 1.37. Adjusting for covariates reduced this figure to 1.19. Psychiatric diagnoses had a stronger association with social assistance entry with a risk ratio of 3.01. Adjusting for covariates reduced this figure to a 2.1-fold risk. Of the specific diagnoses, after adjustments, strong predictors of social assistance entry were also diseases of the nervous system (G00–G99-category adjusted RR=1.33), symptoms, signs and abnormal clinical and laboratory findings, not elsewhere classified (R00–R99 -category adjusted RR=1.29) and injury, poisoning and certain other consequences of external causes (S00–T98 category adjusted RR=1.29).

**Figure 1.**
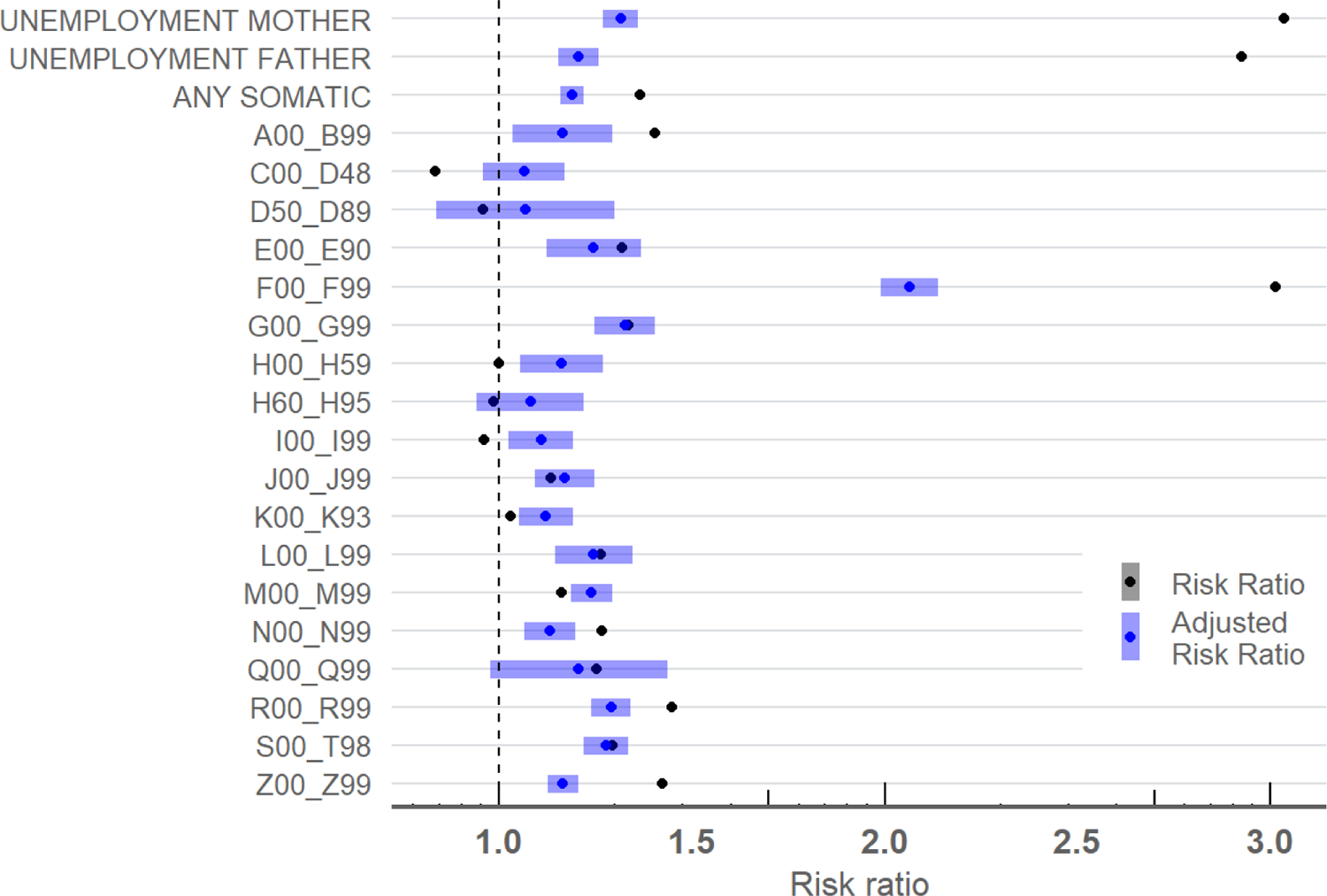
Risk ratios for the associations between different diagnoses associated with a particular medical specialty (ICD-10) and subsequent social assistance entry. Different models were used for all diagnosis groups, with each are estimated separately. Adjusted estimates include 95 percent confidence intervals. Unadjusted n_families= 54 960, n_observations= 801 336, adjusted n_families= 54 843, n_observations = 799 612.

In Figure 2, parental somatic diagnoses had an unadjusted attributable fraction of 12 percent of social assistance entries. Adjusting for covariates reduced this figure to 7.2 percent. Psychiatric diagnoses had an unadjusted PAF of 3.8 percent. Adjusting for covariates reduced this figure to 2.9 percent. Of the specific diagnoses categories, after adjustment, the largest shares of social assistance entries were attributed to factors influencing health status and contact with health services (Z00– Z99 -category adjusted PAF=2.1), symptoms, signs and abnormal clinical and laboratory findings, not elsewhere classified (R00–R99 -category adjusted PAF=2) and musculoskeletal disorders (M00–M99 block, adjusted PAF=1.7).

**Figure 2.**
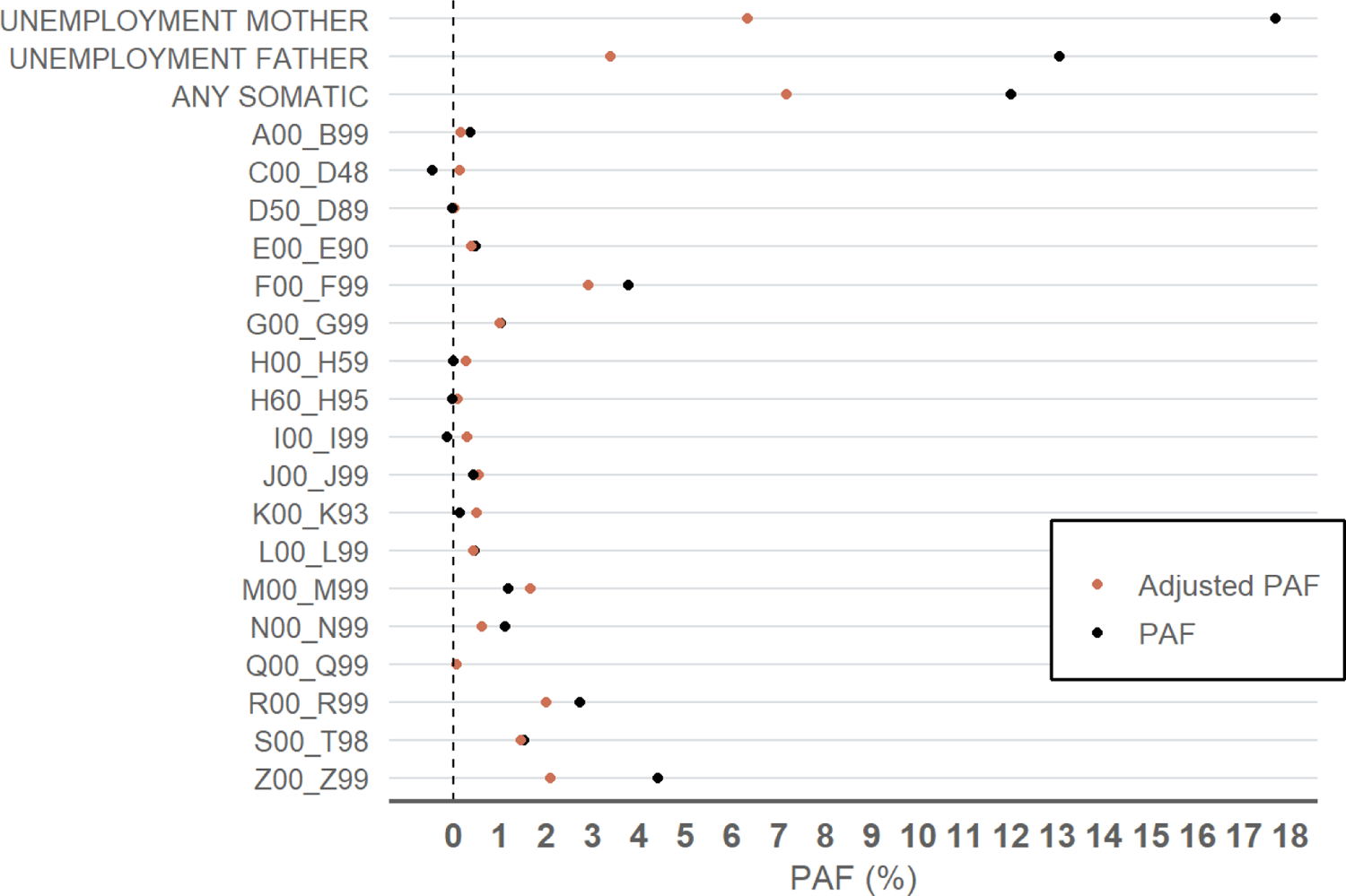
Population attributable fractions for the associations between different psychiatric and somatic diagnoses in specialised care (ICD-10) and subsequent social assistance entry. All diagnosis groups were estimated separately. Unadjusted n_families= 54 960, n_observations= 801 336, adjusted n_families= 54 843, n_observations = 799 612.

We conducted additional analysis by focusing on fathers’ and mothers’ health care records separately in families with two registered parents at the same address (637 867 of observations from 50 171 families). We did not have data on step-parents’ diagnoses. The associations were similar for mothers’ and fathers’ diagnoses with few exceptions. Psychiatric diagnoses had a stronger link to social assistance entry, on a relative scale, when these diagnoses were fathers rather than mothers (Supplementary figure 3 and 4). We also conducted additional analysis for families with a single registered parent living with children. The associations were in a similar direction albeit smaller in relative scale due to a higher share of families with social assistance entry (Supplementary figure 5).

## Discussion

Previous studies have indicated that, in general, health matters for subsequent socioeconomic outcomes and that this is particularly so in respect of social assistance use (11–13,28). Our study is however the first to focus on families with children as the unit of analysis and to provide population-level estimates to illustrate attributable fractions. In addition, the key novelty here is detailed diagnosis-level data on health exposures.

In this study using rich administrative register data, we were able to estimate the contribution of parental health, as proxied by parents’ diagnoses from specialised inpatient and outpatient care records, on the subsequent risk of social assistance entry using rich register data. Our main finding is that parents’ diagnoses, particularly when due to psychiatric conditions, link to a higher risk of social assistance entry, irrespective of parent sex. However, parental somatic and psychiatric diagnoses in total may be attributed to less than a tenth of all social assistance entries.

Our results on the diagnoses category differences indicated that, irrespective of medical specialty, the more the given diagnosis category is concentrated on lower socioeconomic groups the stronger is its link to social assistance use. It is well-established, and certainly unsurprising, that health shocks lead to withdrawal from the labour market and incur some treatment costs. However, in the Finnish context, these types of risks are insured with separate social insurance instruments (29). In fact, the developed welfare net of the country may explain the minimal impact of some diagnoses on social assistance entry.

Data limitations prevent us from exploring the key mechanisms through which diagnoses link to subsequent social assistance entry. Social assistance can be granted to cover medical expenses but it is subject to the same means-testing in respect of assets and income. Nevertheless, our assessment is that treatment expenses alone represent an insufficient explanation for the association observed. Potential mechanisms also include reduced ability to handle finances.

These findings are important in enabling us to recognise target groups and in designing interventions that aim to prevent families with children from falling into economic hardship. The findings presented here suggest that these other social benefits do not fully prevent serious financial difficulties arising from parent ill-health among families with children. Thus, identifying these ‘at risk’ families is a key role when planning preventive actions to avoid the consequences of multiple risk factors in life. These findings also suggest that improving population health by reducing psychiatric disorders in particular, may help to reduce public welfare spending.

The strengths of this study include complete population-level data with very low attrition and no issues with self-reported data. Nevertheless, our findings are limited by the fact that we had data only on registered parents. For example, our data do not have information on the health care records of non-registered parents living with the children. Our findings are however consistent when analyses are restricted to families with two registered parents living with the children. Furthermore, we did not have data on private in-patient and out-patient hospital visits but the share of private hospital usage, at the time of the study, was negligible. Furthermore, the fact that we lacked information on diagnoses in primary care visits implies that we are underestimating the total contribution of parental health problems on social assistance entry. However, primary care visits are typically shorter and may not imply severe health issues, implying that this underestimation may not be substantial. Nevertheless, while there is limited data availability on primary care records, subsequent studies with richer data sources are needed to estimate the total contribution of parental health to socioeconomic disadvantages of the families with children. Finally, some residual confounding is inevitable in the adjusted estimates. The extent of this unmeasured confounding would however have to be substantial in order to fully explain away the key associations.

## Data Availability

Availability of the research data is subject to research permits from the Finnish Institute for Health and Welfare and respective register holders, mandated by Finnish data protection laws and the policies of the register holders.

