## Supplementary materials for "The contribution of parental health to the subsequent social assistance entry of the family with children– a nationwide register-linked birth cohort study in Finland"

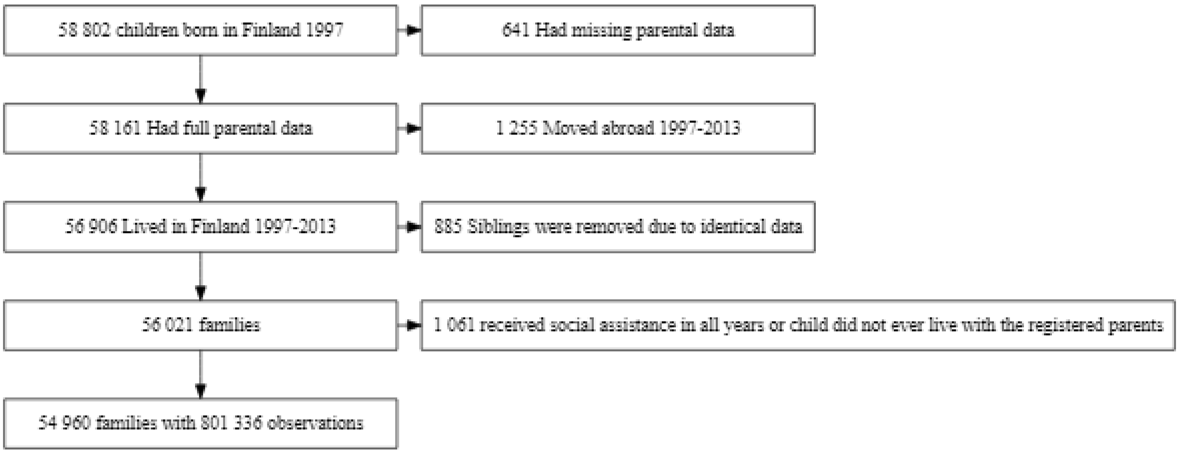


Supplementary figure 1. Description of the study population.

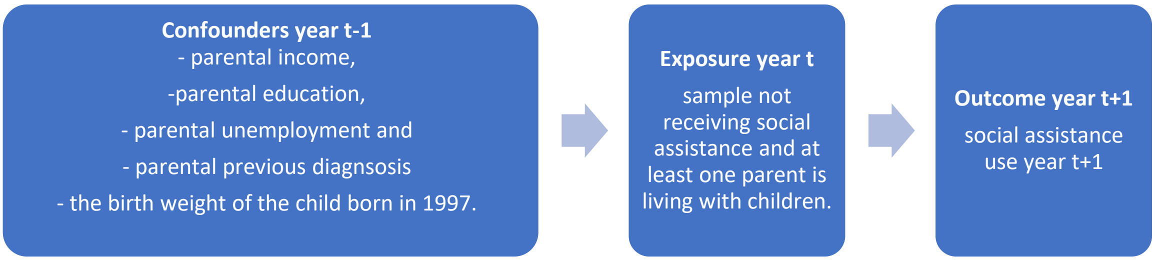


Supplementary figure 2. Description of the study design.

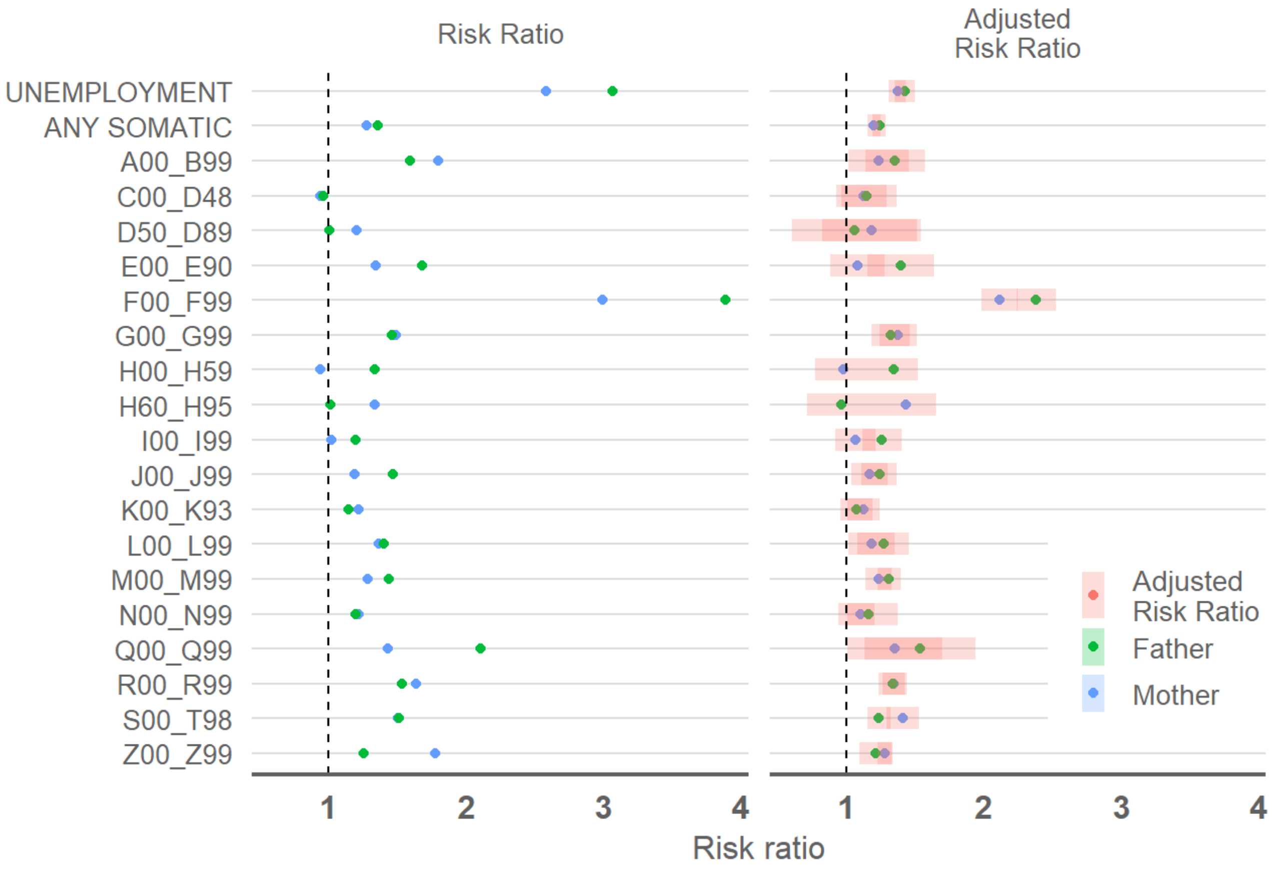


Supplementary figure 3. Risk ratios for the associations between different diagnosis categories from specialised care records (ICD-10) and subsequent social assistance entry during the next year. Different models were used for all diagnose categories, with each estimated separately. Adjusted estimates include 95 percent confidence intervals. The sample is restricted to families with two registered parents living at the same address as the child. Unadjusted n_families= 50 141, n_observations= 637 867, adjusted n_families=50 033, n_observations =  636 503.

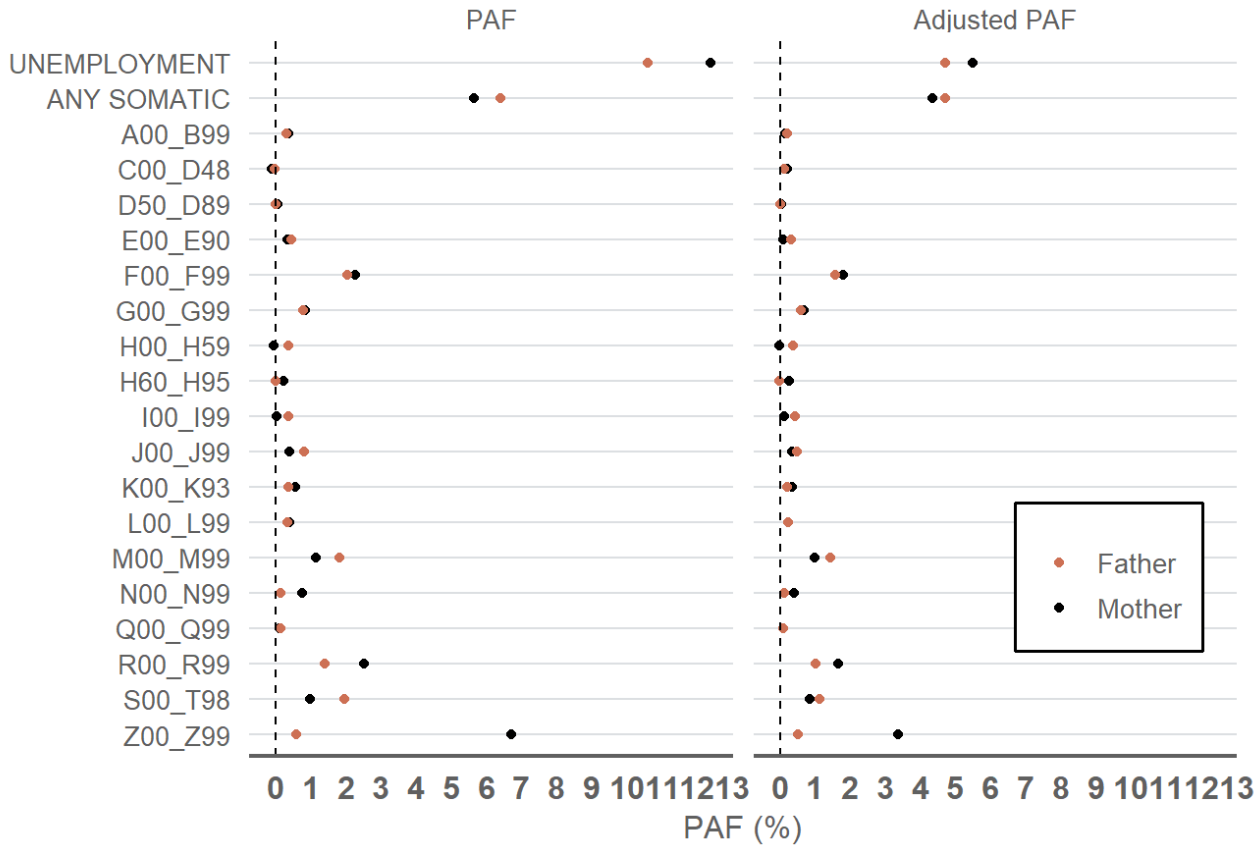


Supplementary Figure 4. Population attributable fractions for the associations between different diagnosis categories from specialised care records (ICD-10) and subsequent social assistance entry during the next year. Different models were used for all diagnose categories, with each estimated separately. Adjusted estimates include 95 percent confidence intervals. The sample is restricted to families with two registered parents living in the same address as the child. Unadjusted n_families= 50 141 , n_observations= 637 867, adjusted n_families=50 033, n_observations =  636 503.

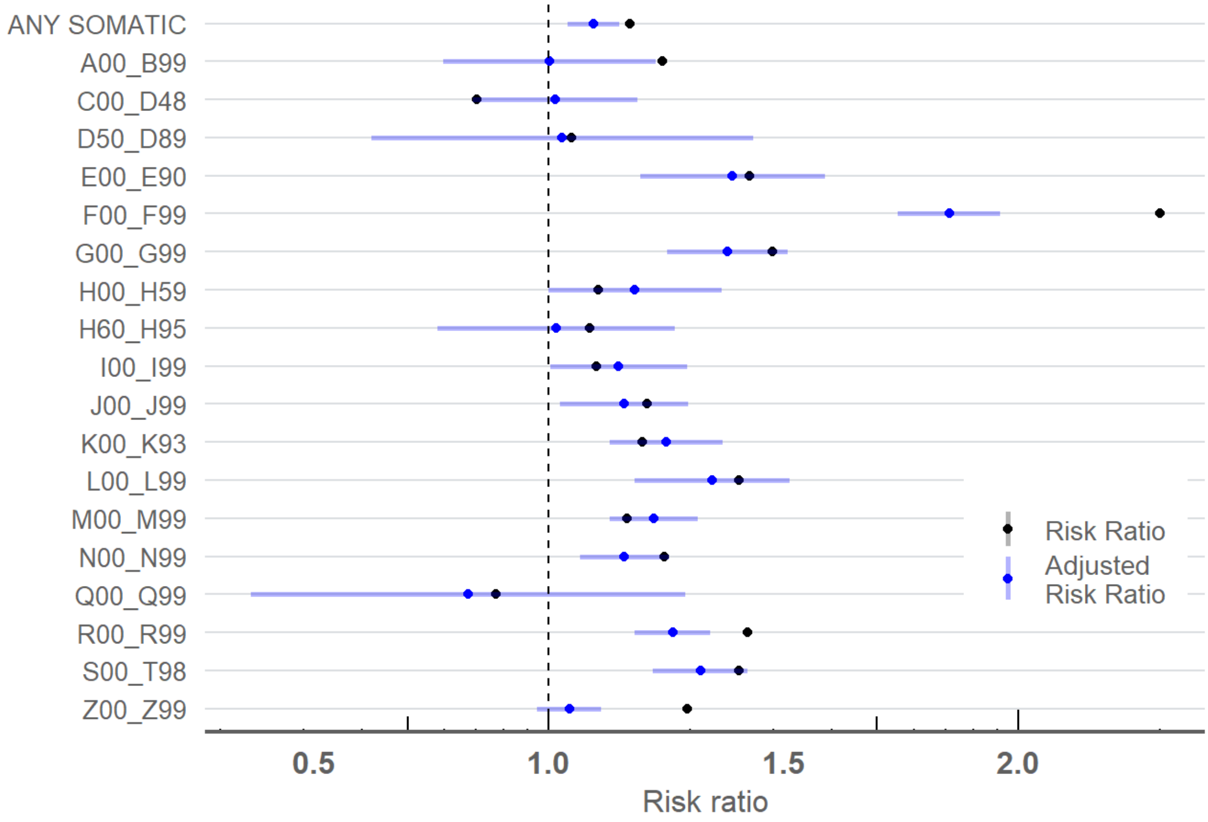


Supplementary Figure 5. Risk ratios fractions for the associations between different diagnosis categories from specialised care records (ICD-10) and subsequent social assistance entry during the next year. Different models were used for all diagnosis categories, with each estimated separately. Adjusted estimates include 95 percent confidence intervals. The sample is restricted to families with one registered parent living in the same address as the child. Unadjusted n_families= 21 022, n_observations= 163 469, adjusted n_families= 20 977 , n_observations = 163 109.

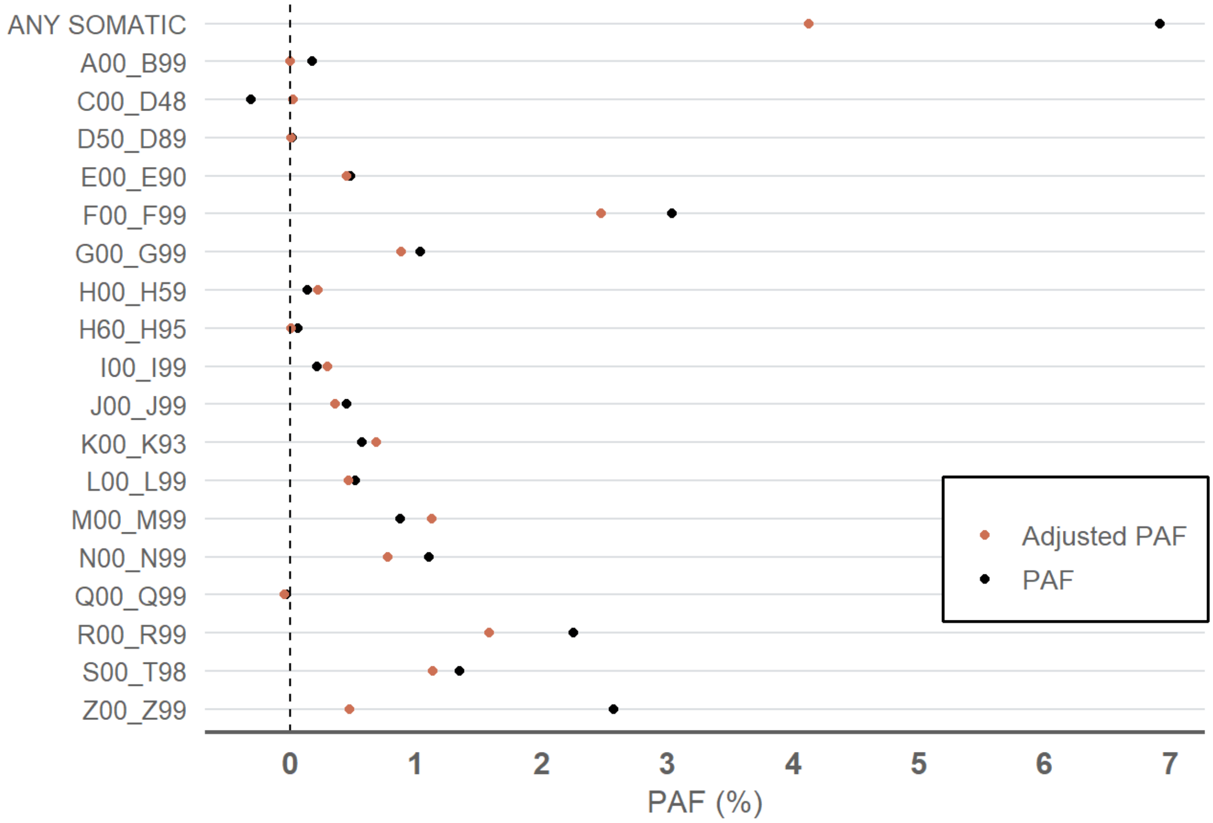


Supplementary figure 6. Population attributable fractions (PAF) for the he associations between different diagnosis categories from specialised care records (ICD-10) and subsequent social assistance entry during the next year. Different models were used for all diagnose categories, with each estimated separately. The sample is restricted to families with one registered parent living in the same address as the child. PAF is calculated using the Miettinen formula. Unadjusted n_families= 21 022, n_observations= 163 469, adjusted n_families= 20 977 , n_observations = 163 109.
